# Global Socioeconomic Context and Brain Ageing in Epilepsy: an ENIGMA-Epilepsy study

**DOI:** 10.64898/2026.04.30.26352177

**Authors:** Heath R Pardoe, Orrin Devinsky, Jemima Robson Bbiomed, Molly Ireland, Julie Absil, Andre Altmann, Marina KM Alvim, Donatello Arienzo, Alice Ballerini, Elisa Barbi, Emanuele Bartolini, Tobias Bauer, Andrea Bernasconi, Neda Bernasconi, Boris Bernhardt, Karen Blackmon, Paolo Bonanni, Leonardo Bonilha, Paolo Bosco, Jacob Bunyamin, Maria Eugenia Caligiuri, Fernando Cendes, Raphaël Christin, Luis Concha, Merran R Courtney, Raul R Cruces, Alberto Danieli, Kathryn A Davis, Chantal Depondt, Patricia Dugan, Gian Marco Duma, John S Duncan, Jerome Engel, Carolina Ferreira Atuesta, Niels KN Focke, Francesco Fortunato, Cesare Gagliardo, Marian Galovic, Antonio Gambardella, Taha Gholipour, Ezequiel Gleichgerrcht, Renzo Guerrini, MV Gule, Ev-Christin Heide, Sara Inati, Victoria Ives-Deliperi, Graeme D Jackson, Sarah Jacobs, Neda Jahanshad, Jonathan K Kleen, Raviteja Kotikalapudi, Mohamad Zakaria Koubeissi, Angelo Labate, Sara Larivère, Matteo Lenge, Helena Martins, Mario Mascalchi, Stefano Meletti, Patrick B Moloney, Terence J O’Brien, Conor Owens-Walton, Costanza Parodi, Antonella Riva, Rebecca W Roth, Jessica Royer, Theodor Rüber, Luca Saba, Ilaria Sammarra, Lucas Scárdua-Silva, Kai Michael Schubert, Leigh Sepeta, Mariasavina Severino, Ben Sinclair, Nishant Sinha, Richard J Staba, Dan J Stein, Joel M Stein, Travis R Stoub, Pasquale Striano, Rainer Surges, Robert Terziev, Sophia I Thomopoulos, Manuela Tondelli, Domenico Tortora, Sebastiano Vacca, Anna Elisabetta Vaudano, Lucy Vivash, Kimberley C Williams, Clarissa L Yasuda, Zhiqiang Zhang, Erik Kaestner, Paul M Thompson, Sanjay M Sisodiya, Carrie R McDonald

## Abstract

**Introduction:** Brain structural differences consistent with an older-appearing brain have been reported in people with epilepsy, but the extent to which these differences reflect clinical characteristics vs broader socioeconomic context is unclear. We investigated whether country-level socioeconomic factors are associated with neuroanatomical differences in adults with epilepsy using MRI-based age prediction, along with epilepsy subtype, sex, and clinical factors.

**Methods:** Structural MRI and clinical data were collected from 26 epilepsy centres across 12 countries in the Americas, Australia, Europe, Asia and Africa. MRI-based age estimates were estimated using a previously developed prediction model trained on 29,175 healthy subjects. Brain predicted age difference (BrainPAD) was calculated as the difference between MRI-predicted brain age and chronological age. National gross domestic product (GDP) per capita and income inequality (Gini index) were obtained from the World Bank. Associations between BrainPAD and epilepsy subtype (temporal lobe epilepsy, extratemporal epilepsy, and genetic generalised epilepsy), national socioeconomic context (GDP per capita and Gini index), age and sex were assessed using regression models.

**Results:** We analyzed 2,109 individuals with epilepsy and 1,041 healthy non-epilepsy controls (57% female; median age = 35; range 17-83). BrainPAD was higher in epilepsy than controls (β 4.2 years, SE 0.4; t=10.6), with increases ranging from 2.5 to 6 years across subtypes. Male sex was associated with 1 year higher BrainPAD relative to females (SE 0.33, t=3.12). There were no main effects of GDP or Gini index; however, significant interactions between were observed. The effect of epilepsy on BrainPAD was greater in countries with lower GDP per capita (t=-2.74) and higher income inequality (t=2.72).

**Conclusions:** Clinical factors and socioeconomic context both influence brain structural ageing in epilepsy. These findings highlight the importance of geographic and economic diversity in neuroimaging research and underscore the relevance of global socioeconomic context when interpreting brain health measures.

## Introduction

Cross-sectional and longitudinal studies show that chronic epilepsy is associated with greater brain atrophy than expected for typical ageing.^1,2^ Machine learning-based techniques can estimate biological age from structural MRI and quantify the difference between MRI-predicted brain age and chronological age, termed brain predicted age difference (BrainPAD). Individuals with treatment-resistant focal epilepsy have been reported to show brains that appear 5-7 years older than expected for their chronological age.^3-5^

Converging evidence from independent studies supports BrainPAD as a proxy measure of brain health. Increased BrainPAD and similar MRI-based metrics are associated with cognitive impairment, subsequent dementia in high-risk individuals, obesity, hypertension and early mortality.^6-11^ Socioeconomic conditions also influence health status and outcomes. Low socioeconomic status is associated with an increased risk for common diseases, including dementia and psychiatric disorders, in large Finnish and UK cohorts.^12^ Neuroimaging studies using functional MRI and EEG have reported associations between biological age and socioeconomic inequality in global cohorts.^13^

We hypothesized that macroeconomic conditions, measured using country-level gross domestic product (GDP) per capita and income inequality (Gini index), may be associated with structural brain ageing in people with epilepsy and healthy controls. To test this hypothesis, we applied an MRI-based age prediction model to structural MRI data from participants in the international ENIGMA-Epilepsy consortium to estimate BrainPAD. The large size and global composition of this cohort enabled us to investigate the relationship between neuroanatomical brain age, disease-related factors such as epilepsy subtype, and national economic conditions. If BrainPAD varies according to socioeconomic context, it may provide a population-level marker of brain health across global settings.

## Methods

### Participants

We analysed structural MRI and associated clinical data collected between 2005 – 2023 through the ENIGMA-Epilepsy consortium. Participating centres were in the United States (10 sites), Australia, Canada, Italy (six sites), Switzerland, Brazil, South Africa, China, Belgium, Germany (two sites), Mexico and the United Kingdom (Figure 1).

**Figure 1.**
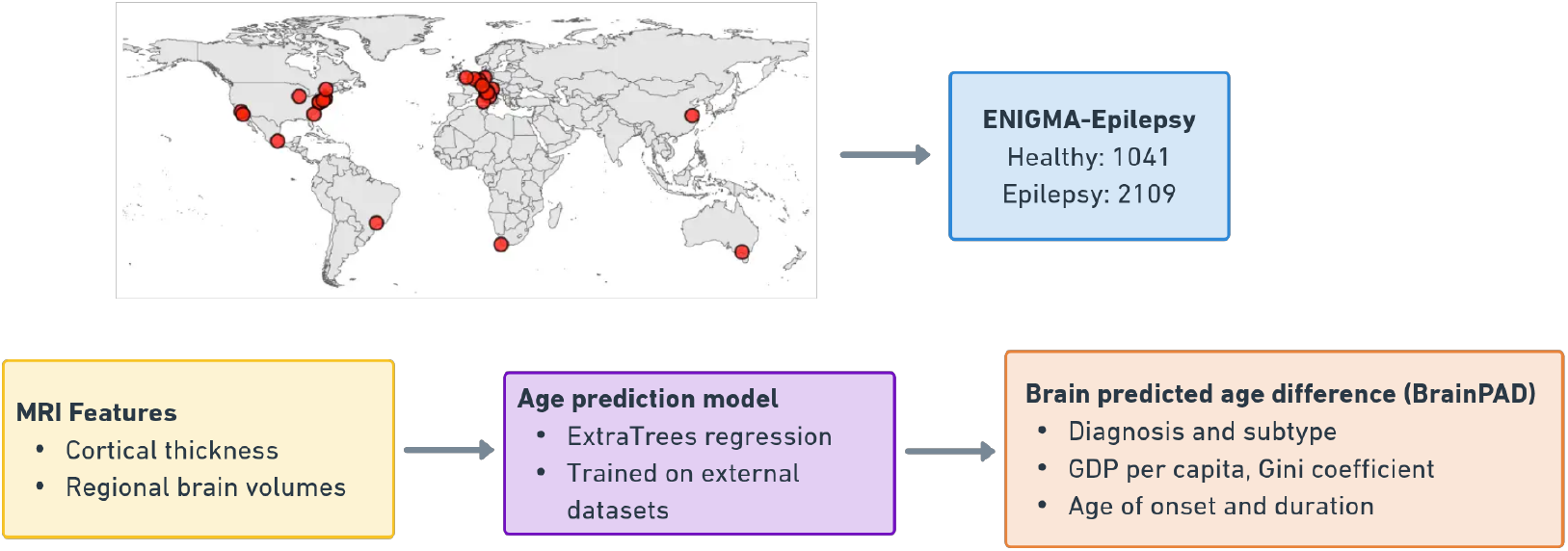
Global distribution of participating ENIGMA-Epilepsy sites and workflow for estimating BrainPAD using structural MRI in the ENIGMA-Epilepsy cohort. Associations between BrainPAD and epilepsy subtype, national-level socioeconomic indicators, and clinical variables were assessed.

**Figure 2.**
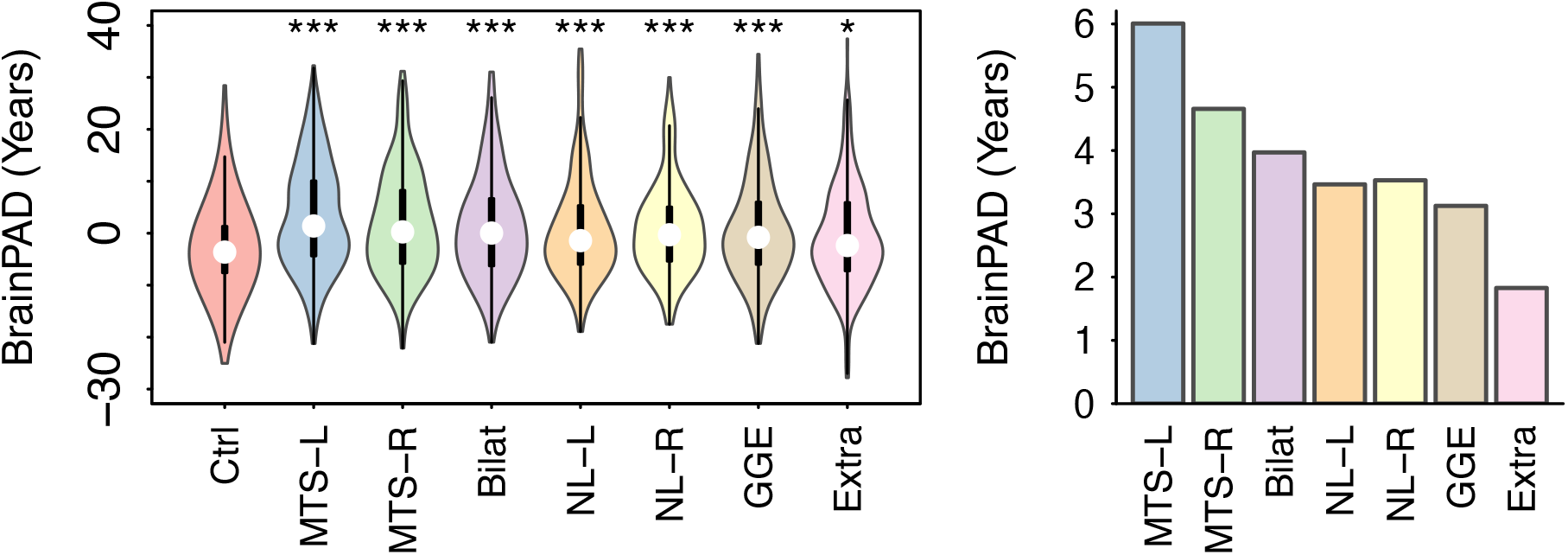
MRI-derived BrainPAD is increased in epilepsy subtypes relative to healthy controls. The left violin plot shows the distribution of BrainPAD values by epilepsy subtype. The right bar chart shows subtype BrainPAD differences after adjustment for covariates. The largest increases in BrainPAD are observed in left and right MTS, whereas lower BrainPAD differences are observed in non-lesional epilepsy cases. Ctrl = Healthy control; MTS-L/R = Mesial temporal lobe epilepsy with hippocampal sclerosis (left or right lateralised); Bilat = bilateral temporal lobe epilepsy; NL-L/R = nonlesional temporal lobe epilepsy left or right lateralised; GGE = genetic generalised epilepsy; Extra = extratemporal non-lesional epilepsy.

Epilepsy participants were included is they had a syndromic diagnosis of epilepsy, had not undergone neurosurgery before imaging, and, for non-lesional groups, had no known abnormalities on clinical MRI (e.g., no large focal cortical dysplasia or other malformations of cortical development, tumors, or other lesions). Healthy controls were recruited locally and age-matched at the site level. Controls were excluded if they had a history of neurological disorders, including epilepsy. Participants with epilepsy and controls were aged 17-83 years.

Phenotypic and clinical data included participant age, sex, epilepsy syndrome diagnosis, age of epilepsy onset, duration of illness and presence of mesial temporal sclerosis (MTS). Seizure and syndromic classification were determined by an epilepsy specialist at each center. Epilepsy syndromes included left and right temporal lobe epilepsy (TLE) with MTS, non-lesional left and right TLE, genetic generalised epilepsy, and non-lesional extratemporal epilepsy.

Country-level GDP per capita and income inequality (Gini coefficient) were obtained from the World Bank’s Open Data portal (https://data.worldbank.org). GDP estimates from 2023 were used. For the Gini coefficient, the most recent available estimate for each country was used. Gini data years ranged from 2014-2022, although estimates from most countries were from 2020-2022.

### MRI acquisition and processing

Structural whole brain T1-weighted MRI scans were collected from each participating center. Images were acquired using 3T scanners (manufacturers Siemens, GE and Philips) with voxel sizes ranging from 0.9 mm isotropic to 1×1×1.3 mm. Further details regarding image acquisition parameters are described in Whelan et al.^14^

Brain age predictions were obtained using a pre-trained brain age prediction model trained on MRI scans of healthy subjects from healthy individuals across multiple repositories, including the UK Biobank (76 sites, https://github.com/james-cole/PyBrainAge^15^). The total number of subjects used for model development = 29,175, 51.5% female, mean age = 46.9 years, SD = 24.4 years. Input features for model training were regional cortical thickness estimates and subcortical and whole brain volume estimates derived using the FreeSurfer software package.^16,17^ Whole brain T1w MRI scans for the ENIGMA-Epilepsy study cohort were processed using Freesurfer to derive regional cortical thickness and brain volume estimates and used to estimate the age of study participants using the age prediction model described above (Figure 1).

Image quality was estimated by calculating the average Euler number for each MRI scan. The Euler number is a topological measure derived from the Freesurfer processing stream that reflects the number of holes or defects in the surface delineating white matter and gray matter. A higher Euler number (less negative) indicates fewer topological defects and better scan quality.^18^

### Statistical analysis

The primary outcome was the BrainPAD, defined as predicted brain age minus chronological age. We fitted a series of linear mixed-effects models with site included as a random intercept to account for clustering within centres. Age and sex were included as covariates in all models. Age was included because brain age prediction models show systematic bias as a function of chronological age, with increased brain age estimates in younger subjects and reduced brain age estimates in older subjects relative to the mean age of the training cohort.^19^

Model 1 assessed the overall association between epilepsy diagnosis (epilepsy vs control) and BrainPAD. Models 2 and 3 examined whether country-level socioeconomic indicators modified this association by including diagnosis-by-GDP per capita and diagnosis-by-Gini interaction terms respectively. Model 4 replaced the binary diagnosis term with epilepsy subtype to assess subtype-specific differences in BrainPAD. Model 5 was limited to the subset of epilepsy cases with age of onset data available (n = 1908, 90% of epilepsy cases) and used least absolute shrinkage and selection operator (LASSO) regression to examine the relative contributions of age of epilepsy onset and epilepsy duration to BrainPAD. Model 6 added Euler number as an index of MRI quality to assess whether image quality influenced BrainPAD estimates.

LASSO regression was used in Model 5 because it performs simultaneous variable selection and regularization, which is advantageous when predictors are highly correlated; in this case age of onset and epilepsy duration are strongly related. The *glmnet* software package was used for LASSO analysis^20^, and the optimal regularisation parameter (λ) was estimated using k-fold cross validation. Linear mixed-effects models were fitted using the lme4 package.^21^ All statistical analyses were performed in R (version 4.4.0^22^).

## Results

We analyzed data from 2,109 individuals with epilepsy and 1,041 healthy non-epilepsy controls (Table 1). Participants with epilepsy had a mean age of 36 years (SD 12), while controls had a mean age of 38 years (SD 12). Females comprised 57% of the epilepsy cohort and 56% of the control group.

**Table 1.** Demographic, clinical, socioeconomic and image quality characteristics of the ENIGMA-Epilepsy dataset.

|  | Epilepsy cases (n = 2109) | Healthy controls (n = 1041) |
| --- | --- | --- |
| Age, mean (SD), years | 35.7 (12.2) | 37.7 (12.3) |
| Sex, n (%) |  |  |
| Female | 1189 (56%) | 595 (57%) |
| Male | 920 (44%) | 446 (43%) |
| MRI quality (Euler number), mean (SD) | -51.2 (76) | -44.3 (108) |
| GDP per capita, \$USD (2023), median [IQR] | 44233 [12908 – 54610] | |
| Gini coefficient, median [IQR] | 35.7 [32.4 – 41.3] |  |
| Age at onset, mean (SD), years | 18 (13.5) | Not applicable |
| Duration of epilepsy, mean (SD), years | 19.5 (14.0) | Not applicable |
| Epilepsy subtype, n (%) |  | Not applicable |
| Genetic generalised epilepsy (GGE) | 267 (13%) | Not applicable |
| Temporal lobe epilepsy (TLE) with mesial temporal sclerosis (left/right) | 448/308 (36%) | Not applicable |
| TLE nonlesional (left/right) | 339/272 (29%) | Not applicable |
| Bilateral temporal lobe epilepsy | 177 (8%) | Not applicable |
| Nonlesional extratemporal epilepsy | 250 (12%) | Not applicable |

BrainPAD was significantly higher in individuals with epilepsy compared with healthy controls (β 4.2 years, SE 0.4, t = 10.6). Across epilepsy subtypes, the largest increases in BrainPAD were observed in temporal lobe epilepsy with mesial temporal sclerosis (MTS), with increases of 6 years for left-sided and 4.6 years for right-sided cases relative to healthy controls. Bilateral temporal lobe epilepsy was associated with an increase of 4.3 years. Non-lesional temporal lobe epilepsy showed more moderate increases in BrainPAD (3.7 years for left-sided, 3.5 years for right-sided), while genetic generalised epilepsy showed a 3.3-year increase. Patients with non-lesional extratemporal epilepsy had a smaller increase in BrainPAD (2.5 years).

GDP per capita across ENIGMA-Epilepsy ranged from $6,022 to $99,564 USD. In mixed-effects models including site as a random intercept, GDP per capita was not associated with BrainPAD overall (t = -0.76). However, a significant interaction between diagnosis and GDP per capita was observed (Figure 3, t = -2.74), indicating that a higher BrainPAD is associated with lower income countries, but only for individuals with epilepsy. When scaled to increments of $10,000 USD, we estimate that BrainPAD is reduced by 0.2 years for every $10k increase in GDP per capita.

**Figure 3.**
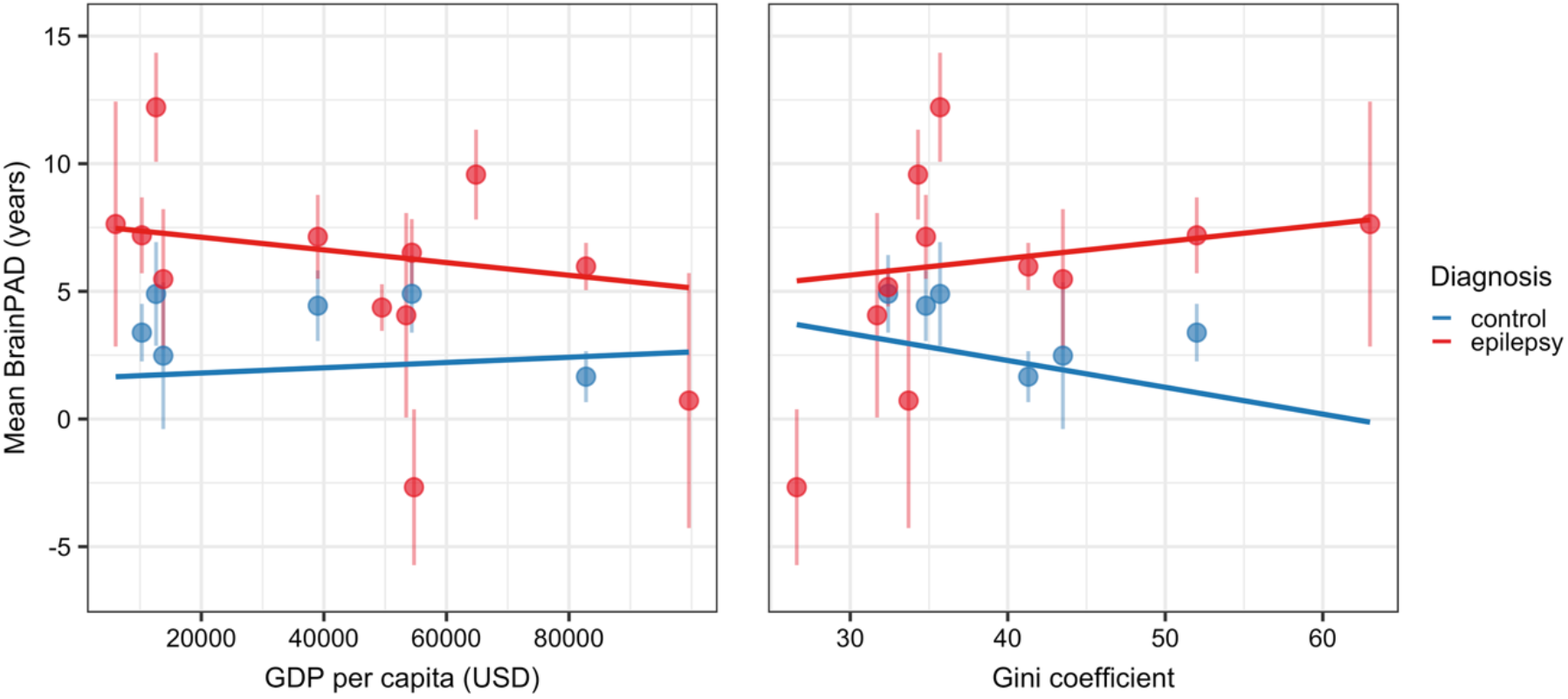
Socioeconomic conditions are associated with structural brain ageing in epilepsy. BrainPAD in people with epilepsy is higher in countries with lower GDP per capita (left), and in countries with greater income inequality (right). Mixed effects models showed significant interactions between diagnosis and socioeconomic measures, indicating that these interactions are primarily driven by epilepsy cases. Points represent country-level mean BrainPAD values, with vertical bars indicating 95% confidence intervals.

Income inequality, assessed using the Gini coefficient, was also not associated with BrainPAD as a main effect (t = -1.29), but a significant interaction between diagnosis and Gini was observed (t = 2.72, Figure 3), indicating higher BrainPAD in epilepsy cases in countries with greater income inequality. An association between BrainPAD and sex was also observed, with females having a BrainPAD one year lower than males (SE 0.33, t=3.12), suggesting comparatively better brain health.

Our LASSO regression analysis revealed that age of onset contributed more to BrainPAD than duration of epilepsy across all epilepsy subtypes. There is a decrease in BrainPAD of 0.6 years for each decade increase in age of onset. MRI image quality, indexed by the Euler number, was comparable between epilepsy cases (mean -55.5, SD 75.7) and controls (mean - 47.3, SD 106.1).

## Discussion

Our analysis of a large, geographically diverse cohort of epilepsy cases and healthy non-epilepsy controls shows that epilepsy was associated with an increased difference between MRI-predicted age and chronological age, ranging from 3.5 years to 5.7 years across epilepsy subtypes. There was no evidence for a main effect of socioeconomic context on BrainPAD, however larger differences between epilepsy and control groups were observed in countries with lower GDP per capita and higher income inequality. These findings suggest that the impact of epilepsy on brain ageing varies according to socioeconomic context, with greater effects observed in more socioeconomically disadvantaged settings.^23^

The observed variation in BrainPAD across socioeconomic settings is consistent with research showing that economic disadvantage is linked to adverse health outcomes across multiple domains including the burden of neurological disease, cognitive function and mental health.^12,24^ Prior work using age prediction models trained on functional MRI and EEG data identified links between increased age gaps and socioeconomic equality in a large international cohort (N = 5,306^13^). That study focused on dementia and included older participants than our cohort (mean age 68 years vs 37 years), which may partly explain differences in findings. For example, they reported an increased age gap in females relative to males, whereas we observed the opposite pattern. However, their finding was limited to females with dementia from Latin American and Caribbean countries; the differences in age, disease-related factors including country-level healthcare access, and the use of functional rather than structural imaging to develop age prediction models may explain this difference. Other studies have reported a reduced brain age gap in females versus males.^7^

With the largest international cohort to date, we demonstrate increased brain ageing effects in epilepsy, consistent with smaller single-site studies.^3-5^ Effect sizes were greater in lesional relative to non-lesional temporal lobe epilepsy cases.^5^ Brain ageing effects in non-lesional extratemporal lobe epilepsy were relatively modest (BrainPAD = 2.5 years); although previous work reported no significant changes in similar cases^5^, our larger cohort was better powered to detect this effect (N = 250 vs 45). Age of epilepsy onset has a stronger association with BrainPAD than epilepsy duration, suggesting that some neuroanatomical differences may be due to developmental or other factors that precede seizure onset. This interpretation is consistent with findings from the Human Epilepsy Project, which identified a relationship between early life learning difficulties and reduced brain volume in adult-onset focal epilepsy.^25^

Limitations of our study include its cross-sectional study design, which precludes causal inferences regarding the association between epilepsy, brain ageing and socioeconomic context. The retrospective nature of our dataset means that image acquisitions were not standardized across epilepsy centers, introducing variability in MRI-based morphometric estimates that could reduce sensitivity to subtle effects.^26^ These limitations are partly offset by the large sample size of the ENIGMA-Epilepsy cohort and the harmonized post-acquisition image processing pipeline used to estimate BrainPAD.

We did not have data on head injury histories or other comorbidities that may influence brain age. Most ENIGMA-Epilepsy sites are associated with tertiary epilepsy care, and our sample is therefore biased towards medically refractory, chronic epilepsy cases. Antiseizure medication use, particularly valproate, was not captured in this study and may contribute to observed brain differences.^27^ Newly diagnosed cases and those that are well-controlled with anti-seizure medication are underrepresented.

The participants included in this study largely come from countries with moderate to high GDP per capita, which may limit the generalisability of our findings across the global socioeconomic landscape. Macroeconomic indicators such as GDP per capita and the Gini index do not capture individual or community-level variability in socioeconomic status.

Future research incorporating detailed individual-level assessments of clinical and family histories, socioeconomic status, and lifestyle factors would provide a better understanding of these relationships.

GDP per capita estimates were derived from 2023 data, and Gini indices from a single year, which did not account for secular changes in economic conditions across participant lifespans. As GDP per capita has generally trended upwards with time in recent decades, younger participants may have been exposed to more favorable economic conditions throughout development and adulthood relative to older subjects. BrainPAD may reflect cumulative lifetime exposures rather than conditions at the time of scanning. These conditions may include access to healthcare, nutrition, education and other determinants of health.

A further limitation is that the brain age prediction model was trained predominantly on data from the USA, United Kingdom and Europe, with more than half of the scans used to train the model from the UK Biobank.^15^ This raises the possibility that systematic differences in model performance across populations may contribute to variation in BrainPAD, particularly in countries or ethnicities underrepresented in the training data. Although we did not observe a main effect of socioeconomic context, the presence of diagnosis-by-socioeconomic interactions suggests that potential bias is unlikely to be uniform across groups. Nevertheless we cannot exclude the possibility that there is an interaction between model performance and disease-related factors. Differences in ethnicity, genetic background, access to healthcare, nutrition and education may influence brain health over the lifespan.^28^ Emerging evidence suggests that large scale environmental stressors, such as climate-related factors, may affect the brain via mechanisms such as heat exposure, pollution, and extreme weather events.^29^ Future research should prioritise the development of brain age prediction models trained on more diverse populations and incorporate a broader set of candidate social, economic and environmental determinants of brain ageing to address these issues. Despite these issues, previous work identified a link between increased BrainPAD and early mortality^7^; the link between country-level wealth and lifespan is well established.^30^

In summary, we demonstrate that adults with epilepsy have older-appearing brains relative to healthy controls, and that the magnitude of this difference varies by clinical factors and country-level socioeconomic context. These findings highlight the importance of geographic and economic diversity in neuroimaging research and underscore the relevance of global socioeconomic context when interpreting brain health measures.

## Supporting information

Supplemental material - author contributions

## Data Availability

Deidentified individual participant data underlying the report findings, including derived brain age estimates (BrainPAD) and associated covariates used in the analyses, will be made available at the time of publication. A data dictionary describing variables will be provided. Additional documents, including the statistical analysis code, will be made available upon reasonable request.
Raw MRI data cannot be shared centrally because of institutional and ethical restrictions across participating sites. Requests for access to raw imaging data should be directed to the corresponding author, who will facilitate contact with individual participating sites, subject to local governance and data sharing agreements.
Data will be made available to qualified researchers for the purposes of replicating findings or conducting secondary analyses, following submission and approval of a research proposal and execution of a data sharing agreement where required. Requests should be directed to the corresponding author.

## Data sharing statement

Deidentified individual participant data underlying the report findings, including derived brain age estimates (BrainPAD) and associated covariates used in the analyses, will be made available at the time of publication. A data dictionary describing variables will be provided. Additional documents, including the statistical analysis code, will be made available upon reasonable request.

Raw MRI data cannot be shared centrally because of institutional and ethical restrictions across participating sites. Requests for access to raw imaging data should be directed to the corresponding author, who will facilitate contact with individual participating sites, subject to local governance and data sharing agreements.

Data will be made available to qualified researchers for the purposes of replicating findings or conducting secondary analyses, following submission and approval of a research proposal and execution of a data sharing agreement where required. Requests should be directed to the corresponding author.

## Acknowledgements

We would like to acknowledge the late Professor Dan J. Stein for his longstanding collaboration within ENIGMA-Epilepsy and for his invaluable contributions to psychiatry, neuroscience, and mental health research. His intellectual generosity, mentorship, and unwavering support shaped this work and the broader field in lasting ways. His scientific legacy and his kindness to colleagues continue to inspire us.

## Notes

### Competing Interest Statement

Paolo Bonanni has received Honoraria from Angelini, Livanova, Jazz, UCB, Neuraxpharm, Eisai.
Niels KN Focke, MD has received Honoraria from Angelini, Jazz Pharma, Bial, Eisai, and Precisis. Research support from Jazz Pharma, all unrelated to this work.
Marian Galovic, MD, PhD has received personal fees and/or travel support from Angelini, Advisis, Bial, Eisai, Neuraxpharm, and UCB.
Antonella Riva has received fees and/or travel support from UCB Pharma, Jazz Pharmaceuticals, BIOCODEX, and STOKE Therapeutics.
Anna Elisabetta Vaudano has received Honoraria from Angelini.

### Funding Statement

ENIGMA extends gratitude to the NIH Big Data to Knowledge (BD2K) award for its foundational support and contributions to consortium development (U54 EB020403 awarded to Paul M Thompson). Research reported in this publication was supported by NIH S10OD032285. For a comprehensive list of ENIGMA-related grant support, please visit: https://enigma.ini.usc.edu/about-2/funding/.
Marina KM Alvim was supported by FAPESP.
Donatello Arienzo, Carrie R. McDonald, Sophia I. Thomopoulos and Paul M Thompson were supported by NINDS R01 NS122827.
Emanuele Bartolini was supported by grant-RC and 5×1000 voluntary contributions, Italian Ministry of Health.
Tobias Bauer was supported by Neuro-aCSis program funded by the Deutsche Forschungsgemeinschaft (DFG, German Research Foundation, 493623632); German Federal Ministry of Research, Technology and Space (epicenter.ai).
Andrea Bernasconi and Neda Bernasconi were supported by Canadian Institute of Health Research.
Karen Blackmon was supported by Wellcome Leap; NIH/NIA (U01AG097744-01); Davos Alzheimer's Association; and U.K. Medical Research Council.
Paolo Bonanni, Alberto Danieli, Gian Marco Duma and Pasquale Striano were supported by Ricerca Corrente 2025 - (Ministero della Salute/Italian Ministry of Health).
Paolo Bosco was supported by Italian Ministry of Health: RC-L4.
Fernando Cendes was supported by FAPESP (Sao Paulo Research Foundation) grant number 2013/07559-3.
Luis Concha was supported by CONACYT (181508, 1782, FC218-2023); and UNAM-DGAPA (IB201712, IG200117, IN204720, IN213423).
Kathryn A Davis was supported by R01-NS-116504.
Orrin Devinsky was supported by Finding A Cure for Epilepsy and Seizures (FACES).
Patricia Dugan was supported by NIH, NeuroPace.
John S Duncan was supported by National Institute for Health and Care Research.
Jerome Engel Jr was supported by Christina Louise George Trust.
Marian Galovic was supported by Swiss National Science Foundation, Swiss League Against Epilepsy, Koetser Foundation for Brain Research, and Herzog-Egli Foundation.
Antonio Gambardella was supported by NEXTGENERATIONEU and funded by the Ministry of University and Research, National Recovery and Resilience Plan, project MNESYS (PE0000006)-A Multiscale Integrated Approach to the Study of the Nervous System in Health and Disease (DN. 1553 11.10.2022).
Taha Gholipour was supported by NIH/NINDS award K23NS135108.
Ezequiel Gleichgerrcht was supported by KL2TR002381& UL1TR002378 Georgia CTSA as well as an American Epilepsy Society (AES) Junior Investigator Award.
Sara Inati was supported by the National Institute of Neurological Disorders and Stroke Intramural Research Program. The content is solely the responsibility of the authors and does not necessarily represent the official views of the National Institutes of Health.
Molly Ireland, Heath Pardoe and Graeme Jackson were supported by the Australian Epilepsy Project which received funding from the Australian Government under the Medical Research Future Fund (Frontier Health and Medical Research Program - Grant Number RFRHPSI000008). Graeme D Jackson was also supported by the Australian Epilepsy Project, funded by the Australian Government Medical; Research Future Fund, Grant/Award Number: MRFF75908.
Sarah Jacobs was supported by the National Research Foundation, (NRF), South African Medical Research Council (SAMRC) and Gabriel Foundation.
Dan J. Stein was supported by the South African Medical Research Council (SAMRC).
Neda Jahanshad was supported by R01AG087513.
Erik Kaestner was supported by NINDS K01NS12483.
Sara Larivere was supported by Centre de recherche du CHUS (CRCHUS), Universite de Sherbrooke, the Natural Sciences and Engineering Research Council of Canada (NSERC: RGPIN-2025-06138), and the Canada Research Chairs program.
Helena Martins was supported by The Amelia Roberts Fund.
Stefano Meletti was supported by the Italian Ministry of Health, NET-2013-02355313.
Terence J O'Brien was supported by NHMRC Investigator Grants (#APP1176426 and APP2034258).
Rebecca Roth was supported by NINDS award T32NS096050.
Jessica Royer was supported by Canadian Institutes of Health Research (CIHR) Fellowship; Natural Sciences and Engineering Research Council of Canada Banting Postdoctoral Fellowship.
Theodor Ruber was supported by the German Ministry of Research, Technology and Research (epi-center.ai).
Kai Michael Schubert was supported by Betty and David Koetser Foundation for Brain Research; Philas Foundation.
Leigh Sepeta was supported by NINDS K23NS093152.
Nishant Sinha was supported by NIH-NINDS K99NS138680.
Sanjay M Sisodiya was supported by Epilepsy Society.
Richard J Staba was supported by R01 NS106957, RF1 NS033310, and Christina Louise George Trust.
Joel M Stein was supported by R01NS116504.
Kimberley C Williams was supported by Carnegie Corporation of New York and the National Research Foundation.
Clarissa L Yasuda was supported by CNPQ(403307/2021-0; 3313263/2025-6; 445340/2024-0).
Renzo Guerrini and Matteo Lenge were supported by the Current Research for IRCCS received from the Italian Ministry of Health in 2025.

### Author Declarations

Ethics committee/IRB of NYU Langone Health gave ethical approval for this work

