## Supplemental material - author contributions for "Global Socioeconomic Context and Brain Ageing in Epilepsy: an ENIGMA-Epilepsy study"

**Title:**

**Contributions**

**Formal analysis**
Heath R Pardoe

**Methodology**
Taha Gholipour, Ezequiel Gleichgerrcht, Molly Ireland, Neda Jahanshad, Heath R Pardoe, Jemima Robson, Paul M Thompson, Manuela Tondelli

**Validation**
Heath R Pardoe, Lucas Scárdua-Silva

**Visualisation**
Heath R Pardoe

**Project administration**
Patricia Dugan, Carrie R McDonald, Terence J O'Brien, Heath R Pardoe, Jemima Robson, Dan J Stein, Sophia I Thomopoulos, Paul M Thompson, Lucy Vivash

**Data curation (imaging and/or clinical)**
Julie Absil, Marina KM Alvim, Elisa Barbi, Tobias Bauer, Karen Blackmon, Leonardo Bonilha, Paolo Bosco, Jacob Bunyamin, Maria Eugenia Caligiuri, Fernando Cendes, Raphaël Christin, Merran R Courtney, Raul R Cruces, Alberto Danieli, Patricia Dugan, John S Duncan, Jerome Engel Jr, Francesco Fortunato, Antonio Gambardella, Taha Gholipour, Ezequiel Gleichgerrcht, MV Gule, Ev-Christin Heide, Molly Ireland, Victoria Ives-Deliperi, Sarah Jacobs, Erik Kaestner, Jonathan K Kleen, Raviteja Kotikalapudi, Matteo Lenge, Mario Mascalchi, Patrick B Moloney, Terence J O'Brien, Conor Owens-Walton, Heath R Pardoe, Costanza Parodi, Ev-Christin Poelstra, Antonella Riva, Rebecca W Roth, Jessica Royer, Theodor Rüber, Luca Saba, Lucas Scárdua-Silva, Kai Michael Schubert, Mariasavina Severino, Ben Sinclair, Nishant Sinha, Richard J Staba, Dan J Stein, Travis R Stoub, Rainer Surges, Manuela Tondelli, Domenico Tortora, Sebastiano Vacca, Anna Elisabetta Vaudano, Lucy Vivash, Kimberley C Williams, Clarissa L Yasuda, Zhiqiang Zhang

**Writing – original draft**
Heath R Pardoe

**Writing – review & editing**
Andre Altmann, Alice Ballerini, Elisa Barbi, Emanuele Bartolini, Tobias Bauer, Andrea Bernasconi, Neda Bernasconi, Karen Blackmon, Paolo Bosco, Maria Eugenia Caligiuri, Fernando Cendes, Raphaël Christin, Luis Concha, Kathryn A Davis, Chantal Depondt, Patricia Dugan, Gian Marco Duma, John S Duncan, Jerome Engel Jr, Carolina Ferreira Atuesta, Niels KN Focke, Cesare Gagliardo, Marian Galovic, Ezequiel Gleichgerrcht, Ev-Christin Heide, Molly Ireland, Victoria Ives-Deliperi, Graeme D Jackson, Neda Jahanshad, Erik Kaestner, Jonathan K

Kleen, Angelo Labate, Sara Larivère, Matteo Lenge, Helena Martins, Mario Mascalchi, Carrie R McDonald, Patrick B Moloney, Terence J O'Brien, Heath R Pardoe, Ev-Christin Poelstra, Antonella Riva, Jemima Robson, Rebecca W Roth, Luca Saba, Mariasavina Severino, Nishant Sinha, Sanjay M Sisodiya, Dan J Stein, Joel M Stein, Pasquale Striano, Rainer Surges, Robert Terziev, Sophia I Thomopoulos, Paul M Thompson, Sebastiano Vacca, Anna Elisabetta Vaudano, Lucy Vivash, Kimberley C Williams, Clarissa L Yasuda

**Formal analysis**
Andre Altmann, Francesco Fortunato, Molly Ireland, Raviteja Kotikalapudi, Heath R Pardoe, Jemima Robson, Manuela Tondelli, Domenico Tortora, Anna Elisabetta Vaudano

**Software**
Donatello Arienzo, Heath R Pardoe, Nishant Sinha

**Cohort PI**
Emanuele Bartolini, Andrea Bernasconi, Neda Bernasconi, Boris Bernhardt, Paolo Bonanni, Fernando Cendes, Luis Concha, Kathryn A Davis, Chantal Depondt, Orrin Devinsky, Patricia Dugan, Gian Marco Duma, Niels KN Focke, Marian Galovic, Antonio Gambardella, Ezequiel Gleichgerrcht, Renzo Guerrini, Sara Inati, Jonathan K Kleen, Mohamad Zakaria Koubeissi, Angelo Labate, Carrie R McDonald, Stefano Meletti, Terence J O'Brien, Theodor Rüber, Luca Saba, Leigh Sepeta, Mariasavina Severino, Nishant Sinha, Sanjay M Sisodiya, Richard J Staba, Pasquale Striano, Zhiqiang Zhang

**Funding acquisition**
Leonardo Bonilha, Fernando Cendes, Kathryn A Davis, Orrin Devinsky, Renzo Guerrini, Carrie R McDonald, Terence J O'Brien, Heath R Pardoe, Richard J Staba, Dan J Stein, Sophia I Thomopoulos, Paul M Thompson

**Supervision**
Fernando Cendes, Patricia Dugan, Francesco Fortunato, Renzo Guerrini, MV Gule, Mario Mascalchi, Terence J O'Brien, Heath R Pardoe, Dan J Stein, Pasquale Striano, Anna Elisabetta Vaudano, Lucy Vivash

**Resources**
Leonardo Bonilha, Maria Eugenia Caligiuri, Orrin Devinsky, Jerome Engel Jr, Niels KN Focke, MV Gule, Graeme D Jackson, Neda Jahanshad, Terence J O'Brien, Heath R Pardoe, Ilaria Sammarra, Lucas Scárdua-Silva, Dan J Stein, Rainer Surges, Paul M Thompson
